# Muscle Strength Explains the Protective Effect of Physical Activity against COVID-19 Hospitalization among Adults aged 50 Years and Older

**DOI:** 10.1101/2021.02.25.21252451

**Authors:** Silvio Maltagliati, Stephen Sieber, Philippe Sarrazin, Stéphane Cullati, Aïna Chalabaev, Grégoire P. Millet, Matthieu P. Boisgontier, Boris Cheval

## Abstract

**Objectives:** Physical activity has been proposed as a protective factor for COVID-19 hospitalization. However, the mechanisms underlying this association are unclear. Here, we examined the association between physical activity and COVID-19 hospitalization and whether this relationship was explained by other risk factors for severe COVID-19.

**Method:** We used data from adults aged 50 years and older from the Survey of Health, Ageing and Retirement in Europe. The outcome was self-reported hospitalization due to COVID-19 measured before August 2020. The main exposure was usual physical activity, self-reported between 2004 and 2017. Data were analyzed using logistic regression models.

**Results:** Among the 3139 participants included in the study (69.3 ± 8.5 years, 1763 women), 266 were tested positive for COVID-19 and 66 were hospitalized. Results showed that individuals who engaged in physical activity more than once a week had lower odds of COVID-19 hospitalization than individuals who hardly ever or never engaged in physical activity (odds ratios = 0.41, 95% confidence interval = 0.22–0.74, *p* = .004). This association between physical activity and COVID-19 hospitalization was explained by muscle strength, but not by other risk factors.

**Conclusion:** These findings suggest that, after 50 years of age, engaging in physical activity more than once a week is associated with lower odds of COVID-19 hospitalization. The protective effect of physical activity on COVID-19 hospitalization is explained by muscle strength.

---

The coronavirus disease 2019 (COVID-19) is an infectious disease caused by the severe acute respiratory syndrome coronavirus 2 (SARS-CoV-2). On 30 January 2020, the COVID-19 outbreak was declared a public international health emergency of international concern, highest level of alarm of the World Health Organization (World Health Organizationa, 2020). To reduce the number of hospitalizations and deaths due to COVID-19, a massive effort has been invested on public health measures (2), pharmacological treatments (3) and vaccines (4). In the meantime, a large amount of research has quickly identified risk factors for severe COVID-19 (5). These risk factors include not only older age, male sex, and underlying chronic conditions (i.e., obesity, cardiovascular disease, lung disease, kidney disease, diabetes, and cancer) (6,7), but also lower physical fitness, as indexed by weaker muscle strength (8) or by lower maximal exercise capacity (9). However, less attention has been paid to behavioral protective factors, which may yet represent modifiable and low-cost levers to support health policies. In particular, physical activity has recently been suggested as a protective factor for severe COVID-19 (10,11,12).

The hypothesized protective effect of physical activity may be explained by at least two pathways. First, physical activity has been associated with a greater functioning of the immune system (13), which may in turn decrease the odds for severe illness following respiratory tracts infections (14). Second, physical activity can affect underlying chronic conditions that have been identified as risk factors for COVID-19 hospitalization. Specifically, studies showed that physical activity reduces the risk of developing several chronic conditions including cardiovascular diseases, type 2 diabetes, cancer, and obesity (e.g., 15), and is associated with higher muscle strength (16). These effects of physical activity on chronic conditions or muscle strength may indirectly reduce the risk for severe COVID-19.

To the best of our knowledge, only a few studies have assessed the association between physical activity and severe COVID-19, as indexed by risks for hospitalization (17,18,19,20). These studies showed that a higher level of physical activity was associated with a lower risk for COVID-19 hospitalization. However, two of these studies were cross-sectional and conducted in a sample of COVID-19 patients (18,20). Moreover, these studies only adjusted for a restricted number of risks factors for COVID-19 hospitalization and did not include muscle strength as a risk factor (17,18,19,20). Finally, these studies did not assess whether the association between physical activity and COVID-19 hospitalization was explained by these risk factors. In sum, the mechanisms underlying the association between physical activity and COVID-19 hospitalization remain unclear.

The objectives of the current study are to test the association between physical activity and the odds of COVID-19 hospitalization, and to investigate whether this association is explained by established risk factors for COVID-19 hospitalization (Figure 1).

**Figure 1.**
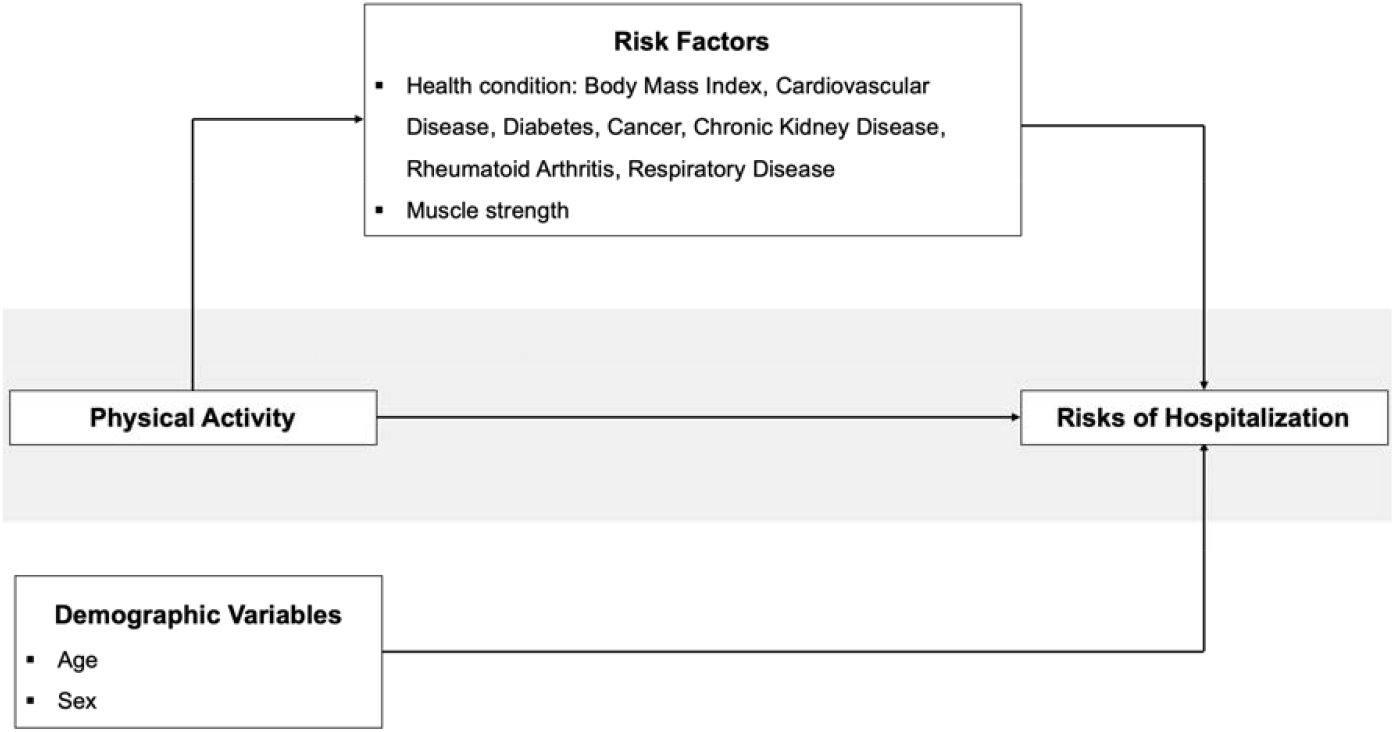
Tested model.

## Methods

### Procedure

SHARE includes longitudinal and cross-national information on socioeconomic circumstances and health from ∼136,000 individuals aged 50 or older living in 27 European countries. Data were collected every two years on seven waves, between 2004 and 2017. In June 2020, a special “SHARE COVID-19” questionnaire was disseminated, assessing social, health, and economic data on ∼52,000 individuals. The present study combines data from these two datasets (i.e., SHARE and SHARE COVID-19). To be included in the current analysis, participants had to be 50 years or older, had completed at least one health questionnaire over the seven survey waves, and had provided an answer to the question “*Have you, or anyone close to you, been tested for the coronavirus and the result was positive, meaning that the person had the COVID disease?*” from the COVID-19 questionnaire. Patients who did not answer this question but indicated that they were hospitalized for COVID-19 were also included in the analysis. SHARE (waves 1-4) was approved by the Ethics Committee of the University of Mannheim. SHARE (waves 4-7) was approved by the Ethics Council of the Max Plank Society.

### Measures

#### Primary Outcome: COVID-19 hospitalizations

COVID-19 Hospitalization was used as an indicator of severe COVID-19 and was determined using the following question: “*Have you or anyone close to you been hospitalized due to an infection from the Corona virus?”* If the patient answered “yes”, the interviewer asked who was hospitalized. Individuals who indicated they were hospitalized were included in the analyses as COVID-19 hospitalized. If the patient indicated that their “*spouse or partner*” was hospitalized, the spouse or partner was included in the analyses as COVID-19 hospitalized.

#### Exposure: Physical activity

Physical activity was assessed using two items, which respectively measured the frequency of low-to-moderate and vigorous physical activity: “*How often do you engage in activities that require a low or moderate level of energy such as gardening, cleaning the car, or doing a walk?” and “How often do you engage in vigorous physical activity, such as sports, heavy housework, or a job that involves physical labor?”* (Cheval et al., 2018). Patients answered on a four-point scale ranging from (*1, More than once a week; 2, Once a week*; *3, One to three times a month; 4; Hardly ever, or never*). Scores were reversed so that so that higher scores reflected higher levels of physical activity. The most recent measure of physical activity over the seven SHARE survey waves was used in the analyses. In the main analysis, the upper value between low-to-moderate and vigorous physical activity was included in the analysis and this variable was treated as a four-level categorical variable. Sensitivity analyses were conducted to examine the distinct association of low-to-moderate and vigorous physical activity with the odds of COVID-19 hospitalization.

#### Other risks factors

We included two covariates related to participants sociodemographic characteristics: *Age* (in 2020, when answering to the SHARE COVID-19 questionnaire), *height*, and *sex* (male, female). Height was adjusted to ensure that the associations between muscle strength and COVID-19 would not simply reflect differences in height (22). We also included eight covariates related to the previous established risk factors for COVID-19 hospitalization: *body mass index* (BMI), *cardiovascular disease* (heart attack, including myocardial infarction or coronary thrombosis, or any other cardiovascular problem including congestive heart failure, high blood cholesterol, high blood pressure or hypertension, stroke or cerebral vascular disease), *diabetes, cancers, chronic kidney disease, rheumatoid arthritis, respiratory disease*, and *muscle strength*.

Muscle strength was indexed by hand grip strength, which was measured using a handheld dynamometer (Smedley, S Dynamometer, TTM, Tokyo, 100 kg) (see 8 for a detailed description of the procedure). The other covariates were measured using self-reported questionnaires. In the case of variables repeatedly assessed across waves, the most recent measures were included in the analyses.

### Statistical analysis

Univariable and multivariable logistic regression models were used to test the association between the exposure of interest (physical activity) and the primary outcome (COVID-19 hospitalization). Model 0 tested the unadjusted association between physical activity and odds of COVID-19 hospitalization. Model 1 tested the associations between physical activity and odds of COVID-19 hospitalization when adjusting for the demographical covariates (i.e., age, height, and sex). These variables were included in Model 1 because they could not explain the potential associations between physical activity and COVID-19 hospitalizations (i.e., physical activity cannot causally predict age, height, or sex). Model 2 added to Model 1 the risk factors (i.e., body mass index, cardiovascular disease, diabetes, cancer, chronic kidney disease, rheumatoid arthritis, respiratory disease) and muscle strength. The *p*-values for global effects were calculated using likelihood ratio tests. To estimate the association of physical activity with COVID-19 hospitalization explained by other risk factors, we computed additional models in which significant risk factors were removed from the equation. In line with previous studies (21, 23), the decrease in the percentage of the association between physical activity and COVID-19 hospitalizations was calculated as follows: 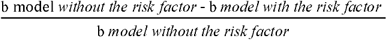, with b representing the estimates of physical activity on COVID-19 hospitalizations. Statistical analyses were conducted in R using the glm package. Statistical assumptions associated with general logistic models were met (i.e., normality of the residuals, multicollinearity, and undue influence).

### Sensitivity analyses

Sensitivity analyses were conducted to 1) examine the distinct association of low-to-moderate and vigorous physical activity with COVID-19 hospitalization.

### Robustness analyses

The dataset was analyzed using a rare-events logistic regression (24), which corrects for the bias associated with rare events. To account for the estimated fraction of patients hospitalized due to COVID-19 in the European population from June to September 2020, we used a tau parameter of 84/100000 based on COVID-19 hospitalization data that were available from May 2020 (see 8 for a detailed description of the procedure). In addition, we corrected for our case-control sampling design using the weighting method of the Zelig package (25).

## Results

The final study sample included 3139 individuals (69.3 ± 8.5 years, 1763 women) (Figure 2), from which 266 were tested positive and 66 were hospitalized for COVID-19 (75,4 ± 10.3 years, 36 women). The number of participants who completed their last measure of usual physical activity in 2004, 2006-2007, 2010-2011, 2012-2013, 2014-2015, and 2017 was 10, 10, 43, 237, 2099, and 740, respectively. Bivariable associations, computed using chi-square tests and correlation tests, showed that physical activity (*p* = .024), higher age (*p <* .001), cardiovascular disease (*p* = .044), and muscle strength (*p* = .003) were associated with COVID-19 hospitalization (vs. no hospitalization). Table 1 summarizes the characteristics of the participants, stratified by COVID-19 hospitalization status.

**Table 1.** Descriptive statistics stratified by hospitalization status.

|  | **Not Hospitalized**<br>(N = 3073) |  | **Hospitalized**<br>(N = 66) |  | **p-values** |
| --- | --- | --- | --- | --- | --- |
|  | N | % | N | % |  |
| **Physical activity** |  |  |  |  |  |
| *Hardly ever or never* | 1131 | 37 | 36 | 54 | .024 |
| *Once to three times a month* | 265 | 9 | 5 | 8 |  |
| *Once a week* | 531 | 17 | 10 | 15 |  |
| *More than once a week* | 1146 | 37 | 15 | 23 |  |
| **Established risk factors** |  |  |  |  |  |
| Age (years) |  |  |  |  |  |
| *50-64* | 1062 | 35 | 9 | 14 | < .001 |
| *65-74* | 1106 | 36 | 21 | 32 |  |
| *75-96* | 905 | 29 | 36 | 54 |  |
| Sex |  |  |  | 0,00 |  |
| *Women* | 1726 | 56 | 37 | 56 | .999 |
| *Men* | 1347 | 44 | 29 | 44 |  |
| Body mass index (kg/m<sup>2</sup>) |  |  |  |  |  |
| *Normal: <25* | 1261 | 41 | 20 | 30 | .142 |
| *Overweight: ≥25 and <30* | 1249 | 31 | 17 | 25 |  |
| *Obese: ≥30* | 563 | 18 | 29 | 44 |  |
| Cardiovascular disease |  |  |  |  |  |
| *No* | 1526 | 50 | 24 | 36 | .044 |
| *Yes* | 1547 | 50 | 42 | 64 |  |
| Respiratory disease |  |  |  |  |  |
| *No* | 2911 | 95 | 59 | 89 | .999 |
| *Yes* | 162 | 5 | 7 | 11 |  |
| Diabetes |  |  |  |  |  |
| *No* | 2782 | 91 | 56 | 85 | .180 |
| *Yes* | 291 | 9 | 10 | 15 |  |
| Cancer |  |  |  |  |  |
| *No* | 2938 | 96 | 63 | 95 | .999 |
| *Yes* | 135 | 4 | 3 | 5 |  |
| Rheumatoid arthritis |  |  |  |  |  |
| *No* | 2825 | 92 | 56 | 84 | .065 |
| *Yes* | 248 | 8 | 10 | 15 |  |
| Chronic kidney disease |  |  |  |  |  |
| *No* | 3033 | 99 | 63 | 95 | .088 |
| *Yes* | 40 | 1 | 3 | 5 |  |
| Muscle strength (kg) (Mean ± SD) | 34.57 ± 11.79 |  | 30.32 ± 10.89 |  | .004 |

*Note:* For muscle strength, mean and standard deviations (SD) are reported. *p*-values are based on the analysis of variance and chi-square tests for continuous and categorical variables, respectively, testing the bivariate association between hospitalization (vs. no hospitalization) and these variables.

**Figure 2.**
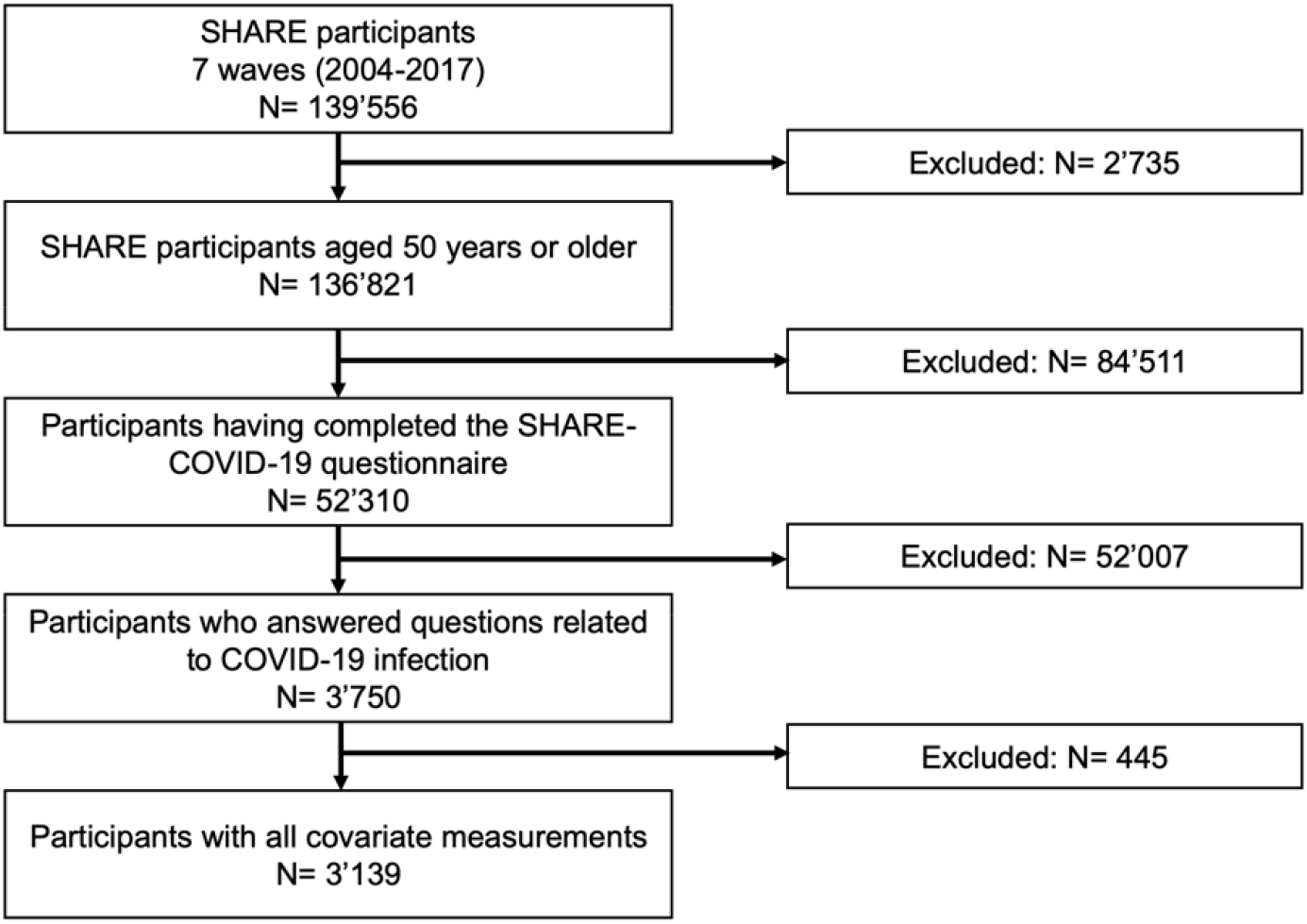
Flow chart of participants.

**Figure 3.**
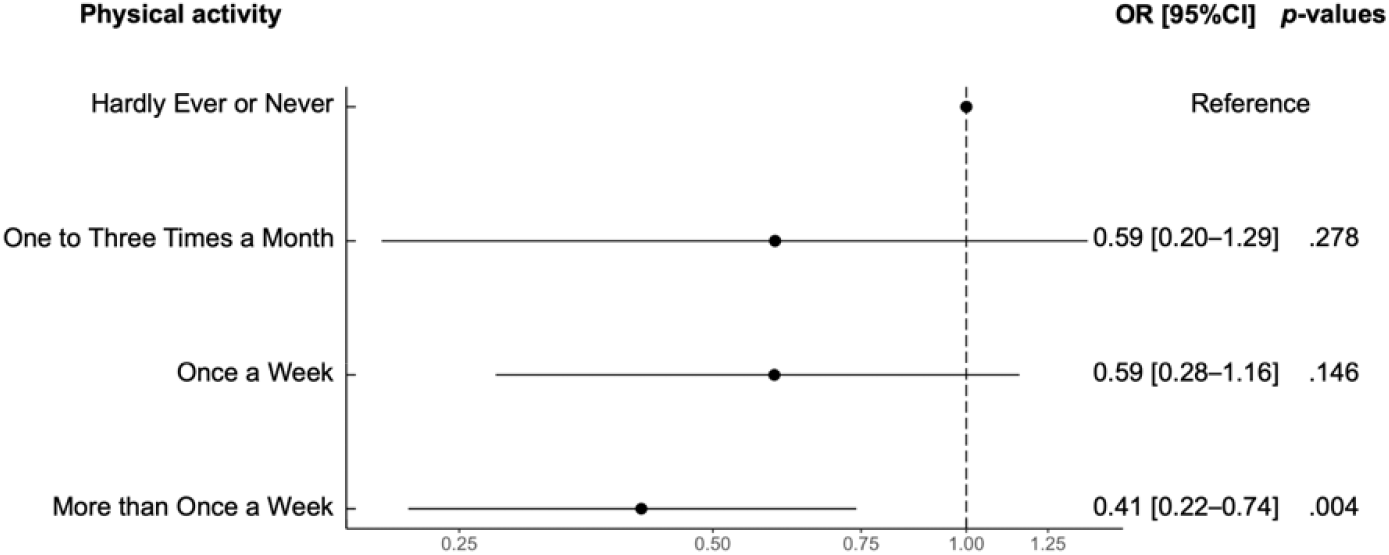
Odds ratios for the different levels of physical activity. *Note:* OR: Odds Ratio; [95%CI]: 95% Confidence Interval. Odds ratios were computed from unadjusted model (Model 0). *Hardly ever or never* served as the reference category.

### Unadjusted association between physical activity and odds of COVID-19 hospitalization (Model 0)

Model 0 showed that physical activity was associated with odds of COVID-19 hospitalization (*p* < .001 for global effect). Compared with participants who *hardly ever or never* engaged in physical activity, the odds of COVID-19 hospitalization were lower for those who engaged in physical activity *more than once a week* (odds ratio [OR] = 0.41, 95% Confidence Interval [95%CI] = 0.22–0.74, *p* = .004). The other comparisons were not statistically significant (*ps* > .146) (Table 2).

**Table 2.**
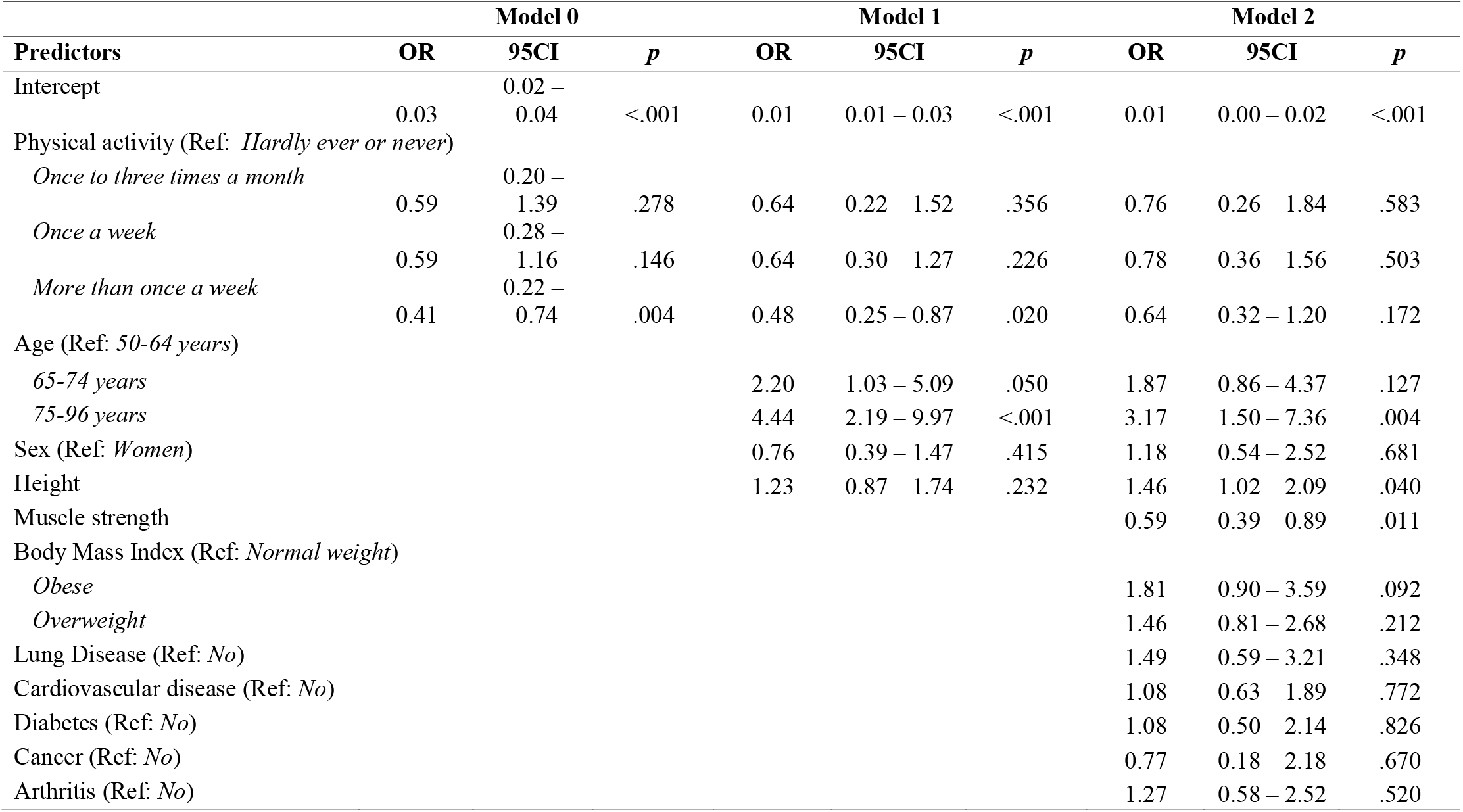
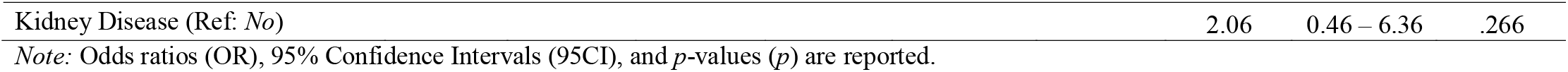
Results of the general logistic models testing the associations of physical activity and other risk factors with COVID-19 hospitalizations.

### Physical activity, demographic variables and odds of COVID-19 hospitalization (Model 1)

Model 1 showed that physical activity remained associated with COVID-19 hospitalization (*p* for global effect = .042). Consistent with the previous model, the odds of COVID-19 hospitalization were lower for participants who engaged in physical activity *more than once a week* than participants who engaged in physical activity *hardly ever or ever* (OR = 0.48, 95%CI = 0.25–0.87, *p* = .020). Moreover, the odds of COVID-19 hospitalization were greater in older participants (*p* < .001 for global effect). Compared to participants aged 50-64 years, participants aged 65-75 years (OR = 2.20, 95%CI = 1.03–5.09, although *p* = .050) and 75-96 years (OR = 4.44, 95%CI = 2.19–9.97, *p* < .001) had greater odds of COVID-19 hospitalization.

### Physical activity, demographic variables, established risks factors and odds of COVID-19 hospitalization (Model 2)

Model 2 showed that physical activity was no longer associated with odds of COVID-19 hospitalization, after adjustment with the other risk factors (*p* = .569 for global effect). Older age remained associated with the odds of COVID-19 hospitalization (*p* for global effect < .001; OR for 75-96 years = 3.17, 95%CI = 1.50–7.36, *p* = .004). Moreover, higher muscle strength was associated with lower odds of COVID-19 hospitalization (OR = 0.59, 95%CI = 0.39–0.89, *p* = .011). In contrast, the other relevant risks factors were not significantly associated with the odds of COVID-19 hospitalization (*ps* > .092). Further analyses revealed that muscle strength explained ∼24% of the association between physical activity and COVID-19 hospitalization.

### Sensitivity analyses

Results of the sensitivity analyses yielded similar results as the main analysis. Specifically, both low-to-moderate and vigorous physical activity were associated with COVID-19 hospitalization in unadjusted models and in models adjusted for demographical variables. Finally, this association was explained by muscle strength (Tables S1 and S2).

### Robustness analyses

The rare-events logistic regression yielded similar results as the main analysis (Table S3).

## Discussion

In this study including 3139 participants, we found that higher physical activity was associated with lower odds of COVID-19 hospitalization. In particular, individuals who usually engaged in physical activity more than once a week had lower odds to be hospitalized due to COVID-19 than those who hardly ever or never engaged in physical activity. Furthermore, after adjustment for a wide range of established risk factors, we found that muscle strength was the only factor explaining this association. This study adds to the growing literature that has hypothesized a link between physical activity and severe forms of. COVID-19 (10,11,12). Crucially, our findings not only support this protective association, but also suggest that muscle strength can underlie it.

Our results are consistent with previous studies observing a negative association between physical activity and the odds of COVID-19 hospitalization (17,18,19,20). This relationship can be explained by the effect of physical activity on the functioning of the immune system (13). As such, physical activity decreases the risk of severe forms of diseases causing respiratory distress (e.g., upper respiratory tracts infections) (14). In the case of COVID-19, it has been suggested that physical activity may reduce the inflammatory response after the infection, thereby exposing individuals to lower the odds of hospitalization (10,11,12). Of note, in our study, we only observed a significant difference between participants who engaged in physical activity *more than once week* vs. those who *hardly ever or never* engaged in physical activity. Yet, this lack of significant difference between the different strata of physical activity may be attributed to the low number of observations in some stratum, thereby generating a large variability in estimated coefficients. For example, although the estimated odds ratios were descriptively in the expected direction (OR = .59 for “*once to three times a month*”, in comparison with “*hardly ever or never*”), only five individuals were hospitalized in the stratum *“once to three times a week”*.

When examining the potential mechanisms underlying the relationship between physical activity and COVID-19 hospitalization, we found that this association was no longer significant, after adjustment for other risks factors. Specifically, our results suggest that muscle strength may play a pivotal role in explaining the protective effect of physical activity on COVID-19 hospitalization. These findings are consistent with previous studies showing that, on the one hand, engaging in regular physical activity reduces sarcopenia (16,26) and that, on the other hand, a greater physical fitness, as indexed by muscle strength, reduced the odds of COVID-19 hospitalization (8). However, to the best of our knowledge, our study was the first to test and demonstrate that the link between physical activity and of COVID-19 hospitalization was explained by muscle strength.

The present study has several strengths, including its prospective design and the adjustment for a wide range of established risk factors for COVID-19 hospitalization. Moreover, the results of the sensitivity and robustness analyses were consistent with the main analysis, strengthening the present findings. However, this work also presents some limitations. First, low-to-moderate and vigorous physical activity were self-reported, which may have reduced measurement validity. Second, because physical activity and other risks factors could have been assessed at a same time, we cannot infer a causal relationship between these variables (i.e., physical activity can affect certain chronic conditions, and vice versa). Third, our study focused on the potential protective role of usual or long-term physical activity patterns but did not investigate how acute level of physical activity at the time of the potential infection can also influence odds of hospitalization. Future studies should determine whether current levels of physical activity are more closely related to risks for COVID-19 hospitalization, than usual physical activity. Fourth, established risk factors for COVID-19 hospitalization were self-reported, which may have decreased the validity of these measures. Similarly, because the latest assessment of these risk factors was at a minimum of two years before patients’ potential infection to COVID-19, some individuals may have contracted the diseases during this period, thereby resulting in a misclassification bias. These latter limitations may explain the non-significant associations between these established risk factors and odds of COVID-19 hospitalization. Future larger-scale studies are needed to examine whether the associations of physical activity with severe COVID-19 may be also explained by the links between physical activity and other relevant chronic conditions (e.g., diabetes, 23).

## Conclusion

This study shows that physical activity is associated with lower odds of COVID-19 hospitalization in adults aged 50 years and older. This association was explained by muscle strength, but not by the other established risk factors for COVID-19 hospitalization. Because of the high prevalence of physical inactivity in the general population (27), especially at older age (21,28,29) and during the COVID-19 pandemic (30,31,32,33), the present findings highlight the need to encourage older adults to regularly engage in physical activity.

## Data Availability

The SHARE dataset is available at http://www.share-project.org/data-access.html.

## Statement of Conflict of Interest and Adherence to Ethical Standards

All authors declare that they have no conflict of interests.

## Author Contributions

All the authors designed the study. S.S. cleaned the data. M.S. analyzed the data. M.S., B.C., M.P.B. drafted the manuscript. All authors critically appraised the manuscript, worked on its content, and approved its submitted version.

## Ethical approval

This study was part of the SHARE study, approved by the relevant research ethics committees in the participating countries

## Inform Consent

All participants provided written informed consent.

## Data sharing

This SHARE dataset is available at http://www.share-project.org/data-access.html.

## Acknowledgements

*This paper uses data from SHARE Waves 1, 2, 3 (SHARELIFE), 4, 5,6, 7 and 8 (DOIs: <u>10..6103/SHARE.w1.600, 10..6103/SHARE.w2.600, 10..6103/SHARE.w3.600, 10..6103/SHARE.w4.600, 10..6103/SHARE.w5.600, 10..6103/SHARE.w6.600, 10.6103/SHARE.w7.711, 10.6103/SHARE.w8cabeta.001</u>*). The SHARE data collection was primarily funded by the European Commission through FP5 (QLK6-CT-2001-00360), FP6 (SHARE-I3: RII-CT-2006-062193, COMPARE: CIT5-CT-2005-028857, SHARELIFE: CIT4-CT-2006-028812) and FP7 (SHARE-PREP: no.211909, SHARE-LEAP: no.227822, SHARE M4: no.261982). Additional funding from the German Ministry of Education and Research, the Max Planck Society for the Advancement of Science, the U.S. National Institute on Aging (U01_AG09740-13S2, P01_AG005842, P01_AG08291, P30_AG12815, R21_AG025169, Y1-AG-4553-01, IAG_BSR06-11, OGHA_04-064, HHSN271201300071C) and from various national funding sources is gratefully acknowledged (see www.share-project.org).

## Supplementary material

**Table S1.**
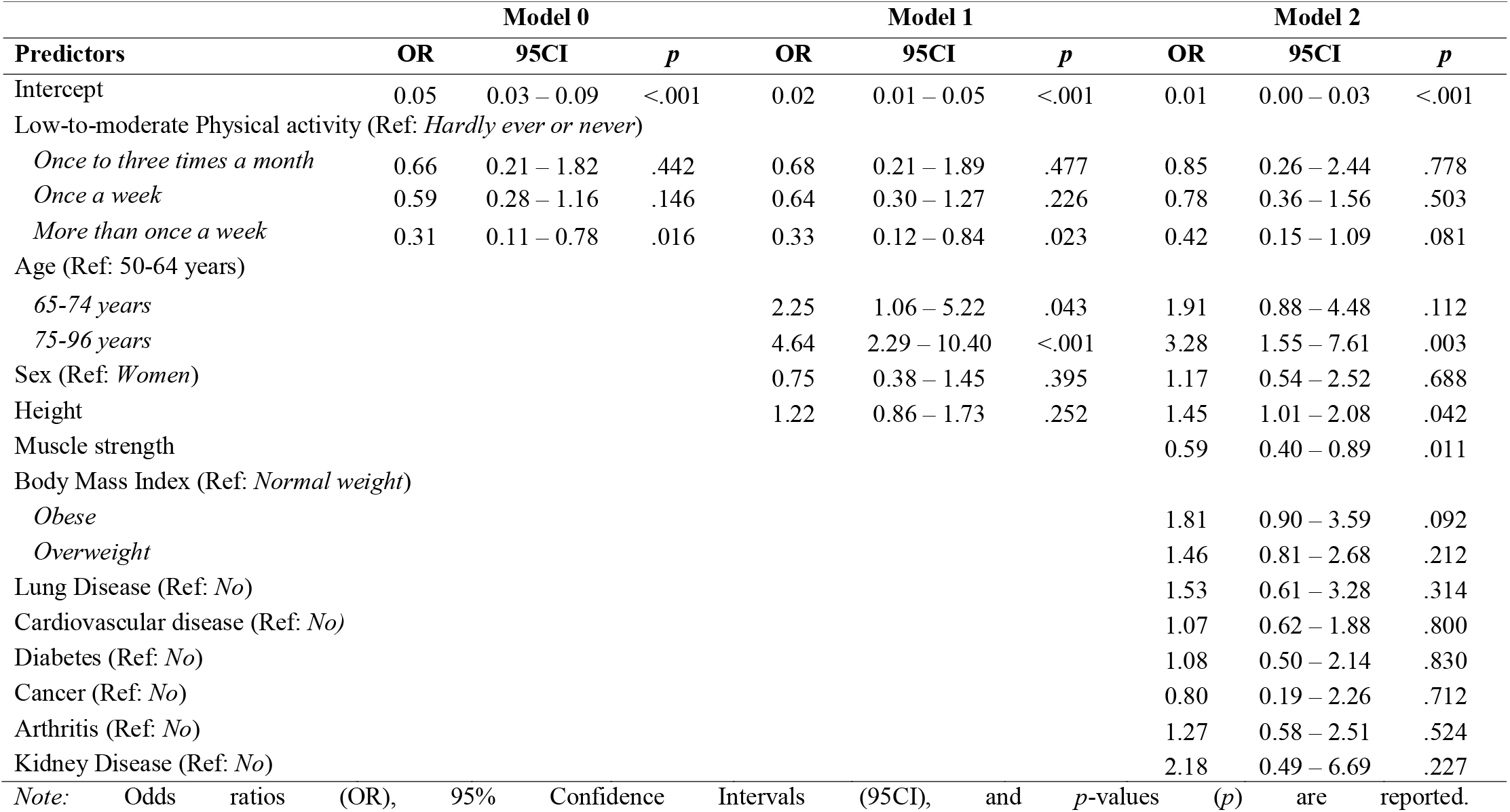
Results of the general logistic models testing the associations of low-to-moderate physical activity and other risk factors with COVID-19 hospitalizations.

**Table S2.** Results of the general logistic models testing the associations of vigorous physical activity and other risk factors with COVID-19 hospitalizations.

| Predictors | Model 0 |  |  | Model 1 |  |  | Model 2 |  |  |
| --- | --- | --- | --- | --- | --- | --- | --- | --- | --- |
|  | OR | 95CI | *p* | OR | 95CI | *p* | OR | 95CI | *p* |
| Intercept | 0.03 | 0.02 – 0.04 | <.001 | 0.01 | 0.01 – 0.03 | <.001 | 0.01 | 0.00 – 0.02 | <.001 |
| Vigorous Physical activity (Ref: *Hardly ever or never*) |  |  |  |  |  |  |  |  |  |
| *Once to three times a month* | 0.66 | 0.22 – 1.55 | .387 | 0.71 | 0.24 – 1.70 | .491 | 0.85 | 0.29 – 2.07 | .751 |
| *Once a week* | 0.65 | 0.30 – 1.28 | .240 | 0.71 | 0.33 – 1.41 | .354 | 0.87 | 0.40 – 1.75 | .711 |
| *More than once a week* | 0.40 | 0.21 – 0.71 | .003 | 0.46 | 0.24 – 0.83 | .012 | 0.61 | 0.31 – 1.13 | .121 |
| Age (Ref: 50-64 years) |  |  |  |  |  |  |  |  |  |
| 65-74 years |  |  |  | 2.20 | 1.03 – 5.09 | .051 | 1.86 | 0.86 – 4.36 | .127 |
| 75-96 years |  |  |  | 4.44 | 2.19 – 9.98 | <.001 | 3.18 | 1.51 – 7.38 | .004 |
| Sex (Ref: *Women*) |  |  |  | 0.77 | 0.39 – 1.48 | .428 | 1.18 | 0.54 – 2.54 | .671 |
| Height |  |  |  | 1.23 | 0.87 – 1.74 | .233 | 1.46 | 1.01 – 2.08 | .041 |
| Muscle strength |  |  |  |  |  |  | 0.59 | 0.39 – 0.89 | .011 |
| Body Mass Index (Ref: *Normal weight*) |  |  |  |  |  |  |  |  |  |
| *Obese* |  |  |  |  |  |  | 1.80 | 0.90 – 3.58 | .093 |
| *Overweight* |  |  |  |  |  |  | 1.45 | 0.81 – 2.66 | .220 |
| Lung Disease (Ref: *No*) |  |  |  |  |  |  | 1.50 | 0.60 – 3.25 | .338 |
| Cardiovascular disease (Ref: *No*) |  |  |  |  |  |  | 1.08 | 0.63 – 1.88 | .777 |
| Diabetes (Ref: *No*) |  |  |  |  |  |  | 1.09 | 0.50 – 2.14 | .822 |
| Cancer (Ref: *No*) |  |  |  |  |  |  | 0.78 | 0.18 – 2.20 | .680 |
| Arthritis (Ref: *No*) |  |  |  |  |  |  | 1.26 | 0.58 – 2.51 | .530 |
| Kidney Disease (Ref: *No*) |  |  |  |  |  |  | 2.07 | 0.47 – 6.42 | .261 |

*Note:* Odds ratios (OR), 95% Confidence Intervals (95CI), and *p*-values (*p*) are reported.

**Table S3.** Results based on the rare-events logistic regression with a tau parameter of 84/100,000.

| Predictors | Model 0 |  |  | Model 1 |  |  | Model 2 |  |  |
| --- | --- | --- | --- | --- | --- | --- | --- | --- | --- |
|  | OR | 95CI | *p* | OR | 95CI | *p* | OR | 95CI | *p* |
| Intercept | 1.3E-2 | 9.1E-4 – 1.7E-3 | <.001 | 5.7E-4 | 2.7E-4 – 1.2E-3 | <.001 | 3.2E-4 | 1.3E-4 – 7.7E-4 | <.001 |
| Physical activity (Ref: *Hardly ever or never*) |  |  |  |  |  |  |  |  |  |
| *Once to three times a month* | 0.65 | 0.25 – 1.66 | .364 | 0.69 | 0.27 – 1.80 | .451 | 0.82 | 0.31 – 2.16 | .695 |
| *Once a week* | 0.61 | 0.30 – 1.25 | .176 | 0.67 | 0.33 – 1.36 | .263 | 0.80 | 0.38 – 1.64 | .550 |
| *More than once a week* | 0.42 | 0.23 – 0.77 | .005 | 0.49 | 0.27 – 0.89 | .019 | 0.65 | 0.35 – 1.22 | .179 |
| Age (Ref: *50-64 years*) |  |  |  |  |  |  |  |  |  |
| *65-74 years* |  |  |  | 2.13 | 0.97 – 4.69 | .060 | 1.81 | 0.81 – 4.03 | .147 |
| *75-96 years* |  |  |  | 4.25 | 2.03 – 8.87 | <.001 | 2.99 | 1.36 – 6.56 | .006 |
| Sex (Ref: *Women*) |  |  |  | 0.77 | 0.37 – 1.60 | .480 | 1.22 | 0.54 – 2.75 | .633 |
| Height |  |  |  | 1.23 | 0.88 – 1.73 | .231 | 1.47 | 1.01 – 2.14 | .047 |
| Muscle strength |  |  |  |  |  |  | 0.57 | 0.37 – 0.89 | .013 |
| Body Mass Index (Ref: *Normal weight*) |  |  |  |  |  |  |  |  |  |
| *Obese* |  |  |  |  |  |  | 1.80 | 0.89 – 3.61 | .101 |
| *Overweight* |  |  |  |  |  |  | 1.47 | 0.82 – 2.66 | .199 |
| Lung Disease (Ref: *No*) |  |  |  |  |  |  | 1.57 | 0.68 – 3.60 | .290 |
| Cardiovascular disease (Ref: *No*) |  |  |  |  |  |  | 1.07 | 0.63 – 1.83 | .802 |
| Diabetes (Ref: *No*) |  |  |  |  |  |  | 1.14 | 0.54 – 2.38 | .735 |
| Cancer (Ref: *No*) |  |  |  |  |  |  | 0.86 | 0.24 – 3.14 | .822 |
| Arthritis (Ref: *No*) |  |  |  |  |  |  | 1.32 | 0.66 – 2.63 | .437 |
| Kidney Disease (Ref: *No*) |  |  |  |  |  |  | 2.55 | 0.73 – 8.83 | .141 |

*Note:* Odds ratios (OR), 95% Confidence Intervals (95CI), and *p*-values (*p*) are reported.

